# Initiation of glucose-lowering drugs reduces the anticoagulant effect of warfarin – but not through altered drug metabolism in patients with type 2 diabetes

**DOI:** 10.1101/2022.10.07.22280782

**Authors:** Ann-Cathrine Dalgård Dunvald, Flemming Nielsen, Dorte Aalund Olsen, Martin Thomsen Ernst, Louise Donnelly, Enrique Soto-Pedre, Maja Refshauge Kristiansen, Jens Steen Nielsen, Frederik Persson, Kurt Højlund, Jonna Skov Madsen, Jens Søndergaard, Ewan Pearson, Anton Pottegård, Tore Bjerregaard Stage

## Abstract

Drug metabolism might be altered in patients with type 2 diabetes. We aimed to evaluate if initiation of glucose-lowering drugs impacts warfarin efficacy and drug metabolism.

First, we conducted a register-based self-controlled cohort study on Danish and Scottish warfarin users. Warfarin efficacy (International Normalized Ratio (INR)) was compared before and after initiation of glucose-lowering drugs. Second, we conducted a clinical pharmacokinetic trial comprising treatment-naïve type 2 diabetes patients. Patients ingested probe drugs for drug-metabolizing enzymes (the Basel Cocktail) before initiating glucose-lowering treatment, and after three and 12 weeks of treatment. Drug metabolism, glycemic control, and inflammation were assessed on each visit.

In the Danish and Scottish cohorts, initiating glucose-lowering drugs reduced warfarin efficacy (n=982 and n=44, respectively). INR decreased from 2.47 to 2.21 in the Danish cohort (mean difference -0.26; 95% CI -0.35;-0.17) and from 2.33 to 2.13 in the Scottish cohort (−0.21; 95% CI - 0.52;0.11) after initiation of glucose-lowering treatment. This impact on INR was more pronounced among individuals with stronger effects of glucose-lowering treatment. In the clinical pharmacokinetic trial (n=10), initiating metformin did not affect drug metabolism after three weeks (geometric mean ratio of CYP3A4 metabolic ratio: 1.12 (95% CI: 0.95;1.32)) or 12 weeks of metformin treatment. Glycemic control improved during treatment, while inflammation remained low and unchanged during treatment.

In conclusion, initiation of glucose-lowering drugs among chronic warfarin users is associated with a reduction in INR, particularly among individuals with a large decrease in HbA_1c_. This effect seems unrelated to CYP enzyme activity and warfarin drug metabolism.

**Registry number:** ClinicalTrials.gov identifier NCT04504045.

## INTRODUCTION

Type 2 diabetes is a chronic metabolic disease caused by a combination of insulin deficiency and insulin resistance and is affecting more than 400 million individuals worldwide (1). Pharmacotherapy is the cornerstone of diabetes management, including glucose-lowering drugs to prevent diabetes-related comorbidities. The most widely used drugs are metabolized by cytochrome P450 enzymes (2), and potential drug-drug interactions are of utmost importance as altered drug metabolism may affect clinical outcomes.

The recent introduction of novel glucose-lowering drugs (3) has changed the landscape of diabetes management. However, metformin remains the first-line treatment for type 2 diabetes (4,5). Metformin is not metabolized by CYP enzymes and is excreted unchanged renally (6). Due to these properties, metformin rarely causes drug-drug interactions (7). However, we and others have previously shown that initiation of metformin caused a decreased efficacy of the anticoagulant vitamin K antagonists warfarin and phenprocoumon (8,9), which are metabolized by CYP enzymes (2). Surprisingly, warfarin’s decreased efficacy was also observed following the initiation of insulin and sulfonylureas (8). The observed effect across drug classes is counterintuitive since these drugs are metabolized and excreted through different pathways and are not expected to affect warfarin metabolism through direct drug-drug interactions. This surprising finding led us to hypothesize that the glucose-lowering effect leads to altered activity of drug-metabolizing CYP enzymes.

Diabetes development and progression are associated with inflammation (10), and inflammation is associated with decreased activity of drug-metabolizing enzymes (11). Patients with type 2 diabetes have reduced activity of drug-metabolizing enzymes compared to non-diabetic individuals (12–14). Still, it is unknown if the activity of CYP enzymes is altered by the initiation of glucose-lowering drugs in patients with type 2 diabetes.

We aim to evaluate the impact of initiating glucose-lowering drugs on drug metabolism. First, we assessed if glucose-lowering therapy affects warfarin efficacy in a register-based study of two cohorts. Second, we aimed to establish the impact of this proposed interaction in a clinical pharmacokinetic trial, comparing the activity of drug-metabolizing enzymes before and after initiation of metformin in treatment-naïve patients with type 2 diabetes.

## METHODS

### Register-based study

#### Data sources and setting

Warfarin users were identified using two independent data sources. The Danish cohort was based on the Copenhagen Primary Care Laboratory (CopLab) database, covering 1.3 million individuals from 2000 to 2015 (15). Data were linked to the Danish Prescription Registry (16) using the unique individual identification number (17). The Scottish cohort was based on laboratory and point-of-care data that covered approximately 400.000 individuals from the areas of Tayside (1992-2021) and Fife (2005-2021). Data were linked to prescription encashment data from Tayside and Fife using the NHS unique individual Community Health Index (CHI) identifier (18).

#### Population

Among warfarin users, we identified individuals with the first prescription of a glucose-lowering drug, defined as the index date. Ongoing long-term treatment was assured by a filled warfarin prescription 180 days before the index date and measurement of INR in the range of eight weeks before to 12 weeks after the index date. We assessed glucose-lowering drugs within each drug class, e.g., insulins, biguanides, sulfonylureas, thiazolidinediones, dipeptidyl peptidase (DPP-4) inhibitors, glucagon-like peptide-1 (GLP-1) analogs, and selective sodium-glucose co-transporter 2 (SGLT2) inhibitors. The Danish cohort did not include users of thiazolidinediones or SGLT-2 inhibitors due to a scarce number of prescriptions during the available study period. We excluded individuals with prescriptions for the same glucose-lowering drug within two years of the index date to ensure initial use. We also excluded individuals initiating another glucose-lowering drug within 120 days of the index date to avoid conflating effects of individual glucose-lowering drugs. Individuals must be over 18 years of age at the index date.

#### Outcome

In a self-controlled design, we compared the INR levels one to four weeks after (short-term effect) and five to seven weeks (long-term effect) after the index date, with the last INR measurement within eight weeks before the index date. We assessed the proportion of individuals with one INR measurement below the therapeutic limit (INR<2) one to six weeks before compared to five weeks after the index date. We conducted several sensitivity analyses in the Scottish cohort. First, we included only individuals at the first-ever prescription of any glucose-lowering drug in the study period. Second, we assessed INR levels in different time ranges before and after the index date. Lastly, we assessed potential confounding by warfarin indication or concomitant use of potential inhibitors (amiodaron, fluconazole, erythromycin, ciprofloxacin, and metronidazole).

We assessed the relationship between INR difference and the last glycated hemoglobin (HbA_1c_) measurement within one year of the index date. Pre-index HbA_1c_ levels were categorized into four groups: < 48 mmol/mol, 48-57 mmol/mol, 58-75 mmol/mol, and >75 mmol/mol. Furthermore, we assessed the change in HbA_1c_ following the initiation of a glucose-lowering drug; we compared the last measurement within one year of the index date with the first measurement within two to six months after the index date. The change in HbA_1c_ was categorized into three groups: an increase (difference ≥ 0), a small decrease (0 to 10 mmol/mol), and a large decrease (more than 10 mmol/mol).

#### Ethics

The Danish cohort was exempt from ethical approval according to Danish law. The Scottish cohort was covered by general ethics and data protection approvals for anonymized record linkage studies, which were obtained for projects hosted by Dundee Health Informatics Center.

#### Statistics

Differences in INR were tested by paired t-test and non-parametric Wilcoxon signed-rank test for small samples. Only results with n>5 are shown to limit random effects by low power.

### Clinical pharmacokinetic trial

We conducted a self-controlled, clinical pharmacokinetic trial to assess the impact of initiating metformin treatment on the activity of drug-metabolizing enzymes in patients with treatment-naïve type 2 diabetes.

#### Trial participants

Patients with type 2 diabetes were recruited at multiple general practices across the Region of Funen, Denmark, from December 2020 to May 2022. Eligible patients were aged 18-75 years with a body mass index (BMI) ≤ 40 kg/m^2^ and no current use of glucose-lowering treatment. Individuals were excluded if they suffered from self-reported inflammatory diseases or cancer, had excessive alcohol consumption defined according to national guidelines, or took medication deemed to affect the safety of the individual or the outcome of the trial. A blood sample was conducted before inclusion to confirm HbA_1c_ ≥ 48 mmol/mol and Glutamic Acid Decarboxylase (GAD-65) antibody, liver-, and kidney function within normal range.

#### Trial medication and dose

The patients self-administered 500 mg metformin twice daily on days 2-8. On day nine and the remaining trial period, the administered dose was increased to 1000 mg metformin twice daily (**Figure S1**). Both amounts are used in routine clinical practice. Compliance was assessed by interviewing and counting tablets, and at least 70% compliance was required to be included in the data analysis. The Basel cocktail was administered orally on the three trial days and consisted of 100 mg caffeine, 50 mg efavirenz, 12.5 mg losartan, 10 mg omeprazole, 12.5 mg metoprolol, and 2 mg midazolam. Complete manufacturing details in **Supplementary**.

#### Trial days

Patients participated in three trial days: before metformin (baseline, day 1), after 3 weeks of metformin treatment (day 22 ± three days), and after 12 weeks of metformin treatment (day 85 ± three days) (**Figure S1)**. The patients fasted, except for water, for 12 hours before the trial days, and fasting was continued until 3 hours after ingestion of the Basel cocktail. In addition, 48 hours before each trial day, the patients were restricted from consuming bitter oranges, grapefruit, alcohol, caffeine, and theobromine. The Basel cocktail was administered with 75 g glucose in an oral solution to perform an oral glucose tolerance test (OGTT). Blood samples were collected at 0 (before administration of the Basel cocktail), 0.5, 1, 1.5, 2, 4, and 6 hours (**Figure S1**). Urine was collected from 0 to 6 hours.

#### Trial approvals and registrations

The trial was approved by the Regional Scientific Ethics Committee of Southern Denmark (identifier S-20200014), the Danish Medicines Agency (identifier 2019111719), and registered in the EudraCT database (identifier 2020-000162-42) and ClinicalTrials.gov database (identifier NCT04504045). The trial was conducted by the Helsinki Declaration and Good Clinical Practice (GCP) and monitored by the GCP unit at Odense University Hospital. Trial subjects were included following written, informed consent.

#### Analytical methods

Analyses for drug and metabolite concentrations were conducted at University of Southern Denmark. All samples were analyzed at the end of the trial and blinded to clinical data. Drugs and metabolites were analyzed in EDTA plasma and urine after a sample pretreatment procedure with enzyme deglucuronidation followed by protein precipitation with standard internal solutions. We used high-performance liquid chromatography (LC) and high-resolution mass spectrometry (HR-MS) following an approach previously described with minor modifications to fit HR-MS (19–22). We used quality control (QC) samples, blanks, and calibration curves in each batch to assess precision and accuracy. The within-batch and between-batch precision (coefficient of variation (CV)) and accuracy (bias) were <15% for all compounds. The calibration curves were linear with r^2^>0.99. The limit of detection (LOD) ranged from 0.1 to 1 ng/mL, and the limit of quantification (LOQ) ranged from 0.5 to 5 ng/mL (**Supplementary**).

Glucose and HbA_1c_ were analyzed continuously during the trial as a part of the daily routine at the Odense University Hospital. Samples were analyzed using Cobas 8000 Roche and TOSOH G8, Alere, respectively. Insulin and C-peptide were analyzed at the end of the trial using Cobas E411.

Analyses for hsCRP, IL-6, IL-1β, TNF-α, and IFN-γ were performed at Lillebælt Hospital. hsCRP was analyzed using Roche/Hitachi cobas c701/702. IL-6 was measured using Cobas e 801 Commercially available kits for the Simoa HD-X analyzer (Quanterix©, Billerica, MA, USA) were used to quantify IL-1β, TNF-α, and IFN-γ in plasma according to the manufacturer’s procedure. Plasma samples were analyzed blinded to clinical data. Quality control was performed using two controls prepared from commercially available control material provided by the manufacturer. The analytical variations were calculated to be <16 % in the IL-1β assay, <13 % in the TNF-α assay, and <6% in the INF-γ.

Genotyping was conducted using a previously validated 5’ nuclease real-time polymerase chain reaction (PCR) test panel (23) to identify the most common *CYP2C9* alleles (*1, *2, *3), *CYP2C19* alleles (*1, *2, *3, *4, and *17) and *CYP2D6* alleles (*1, *2, *3, *4, *5, *6, *9, *10, *17, and *41 or gene duplicates).

#### Statistics and pharmacokinetic analyses

We aimed to include 12 patients in the trial to detect a ≥35% change in midazolam metabolic ratio with 80% power, a two-sided significance level of 5%, and allowing for a 20% drop-out. Recruitment was delayed due to the COVID-19 pandemic, and the trial prematurely ended on May 5^th^, 2022, following the completion of trial subject number ten. Statistical and pharmacokinetic analyses were conducted as previously described (24). In brief, demographic data and pharmacokinetic parameters are shown as medians with interquartile ranges (IQR; 25^th^-75^th^ percentiles) or geometric mean ratios (GMR) with 95% confidence intervals. Non-compartmental analysis was computed using the R package PKNCA (25). AUC_inf_ is only presented when the AUC percent extrapolated is below 25%. Post hoc analysis of efavirenz elimination half-life was calculated with non-compartmental analysis using the concentration from 0 hours at three weeks as a data point for the baseline analysis and time-after-dose was calculated based on days between visits. In the primary analysis, the metabolic ratio was calculated as [drug]/[metabolite] at the time points previously shown to have the highest correlation to the AUC ratio (26). The formation clearances (CL_f_) were estimated as [amount of metabolite in urine_0-6h_] over [AUC of substate_0-6h_], and renal clearances (CL_R_) were calculated as [amount of substrate in urine_0-6h_] over [AUC of substate_0-6h_]. AUC was determined for glucose, insulin, and C-peptide using the same methods as the pharmacokinetic parameters (24).

To assess the random effects of baseline HbA_1c_ and change in HbA_1c_, we used a longitudinal mixed-effect model with the metabolic ratio as the outcome and a fixed interaction term between the variable and time. Change in HbA_1c_ was grouped in the same ranges as used in the register-based study.

## RESULTS

### Register-based study

The Danish and Scottish cohorts included 1,026 individuals (982 and 44, respectively). The median age was 73 years (interquartile range (IQR): 65-80) and 74 years (IQR: 67-79), respectively. Males comprised 56% of the Danish and 52% of the Scottish cohorts.

In the Danish cohort, initiation of any glucose-lowering drug caused a mean change in INR of -0.22 (95% CI: -0.30; -0.13) one to four weeks after the index date. A similar change was observed five to seven weeks after the index date (**Table 1**). Analysis of the individual drug classes showed similar effects for insulin, metformin, and sulfonylureas, while data on DPP-4 inhibitors and GLP-1 receptor analogs did not affect warfarin efficacy (**Table 1**). Data from the Scottish cohort showed no effect for all glucose-lowering drugs one to four weeks after the index date (**Table 1**). Five to seven weeks after the index date, the mean INR changed by -0.21 (95% CI: -0.52; -0.11). A sensitivity analysis, including only the first-ever prescription of any glucose-lowering drug in the Scottish cohort, provided comparable estimates for metformin between the two cohorts (mean INR difference -0.18 (95% CI: -0.69;0.33; n=13) one to four weeks after the index date and -0.32 (95% CI: -0.84; 0.20; n=14) five to seven weeks after the index date). The remaining sensitivity analyses did not yield different results than the main estimates (data not shown).

**Table 1.** The anticoagulant efficacy of warfarin is reduced among patients initiating glucose-lowering treatment in the Danish and Scottish cohorts. Warfarin efficacy was assessed by the international normalized ratio (INR) in patients treated with warfarin before and after initiation of glucose-lowering treatment. In the Danish cohort, more patients experience INR below the therapeutic range (INR< 2) 1-3 weeks after initiating glucose-lowering treatment compared to 2-4 weeks before.

|  | Danish cohort |  |  | Scottish Cohort |  |  |
| --- | --- | --- | --- | --- | --- | --- |
|  | n | Mean INR<br>Before/After | Mean difference<br>(95% CI) | n | Mean INR<br>Before/After | Mean difference<br>(95% CI) |
| **All glucose-lowering drugs** |  |  |  |  |  |  |
| 1-3 weeks after | 677 | 2.49 / 2.27 | -0.22 (-0.30; -0.13) | 32 | 2.55 / 2.50 | -0.05 (-0.36; 0.26) |
| 5-7 weeks after | 539 | 2.47 / 2.21 | -0.26 (-0.35; -0.17) | 31 | 2.33 / 2.13 | -0.21 (-0.52; 0.11) |
| INR < 2 | 540 | 28.3%/39.3% | - | 30 | 45.5%/43.3% | - |
| **Insulin** |  |  |  |  |  |  |
| 1-3 weeks after | 192 | 2.44 / 2.08 | -0.36 (-0.53; -0.19) | 17 | 2.52 / 2.96 | 0.45 (-0.69; 1.58) |
| 5-7 weeks after | 156 | 2.44 / 2.26 | -0.18 (-0.39; 0.02) | 15 | 2.66 / 2.76 | 0.11 (-0.58; 0.79) |
| INR < 2 | 170 | 41.8%/55.3% | - | 16 | 66.7%/25.0% | - |
| **Metformin** |  |  |  |  |  |  |
| 1-3 weeks after | 626 | 2.48 / 2.30 | -0.18 (-0.26; -0.09) | 24 | 2.38 / 2.50 | 0.12 (-0.25; 0.49) |
| 5-7 weeks after | 523 | 2.42 / 2.20 | -0.22 (-0.30; -0.14) | 21 | 2.32 / 2.25 | -0.08 (-0.46; 0.31) |
| INR < 2 | 496 | 28.5%/36.1% | - | 22 | 39.1%/36.4% | - |
| **Sulphonylureas** |  |  |  |  |  |  |
| 1-3 weeks after | 329 | 2.45 / 2.28 | -0.17 (-0.28; -0.06) | 23 | 2.62 / 2.72 | 0.10 (-0.28; 0.48) |
| 5-7 weeks after | 244 | 2.45 / 2.27 | -0.18 (-0.31; -0.05) | 22 | 2.41 / 2.51 | 0.12 (-0.50; 0.74) |
| INR < 2 | 267 | 29.9%/41.9% | - | 21 | 40.9%/47.6% | - |
| **DPP-4 inhibitor** |  |  |  |  |  |  |
| 1-3 weeks after | 138 | 2.38 / 2.31 | -0.08 (-0.28; 0.12) | - | - | - |
| 5-7 weeks after | 103 | 2.39 / 2.42 | 0.03 (-0.20; 0.26) | - | - | - |
| INR < 2 | 113 | 34.9%/31.9% | - | - | - | - |
| **GLP-1 receptor agonist** |  |  |  |  |  |  |
| 1-3 weeks after | 32 | 2.26 / 2.32 | 0.06 (-0.11; 0.22) | - | - | - |
| 5-7 weeks after | 26 | 2.37 / 2.72 | 0.35 (-0.02; 0.72) | - | - | - |
| INR < 2 | 30 | 17.2%/30.0% | - | - | - | - |

Abbreviations: CI: confidence interval, DPP-4: dipeptidyl peptidase-4, GLP-1: Glucagon Like Peptide 1, INR: international normalized ratio, n: number

Baseline HbA_1c_ did not affect the difference in INR levels following the initiation of glucose-lowering treatment (**Figures 1a** and **1c**). However, the change in HbA_1c_ levels after initiation of glucose-lowering therapy compared to baseline is associated with the degree of INR reduction. Patients with a large decrease in HbA_1c_ (more than 10 mmol/mol) had a more pronounced reduction in INR, mean difference -0.44 (95% CI -0.71;-0.18) and -0.40 (95% CI -1.06;0.26), in the Danish and Scottish cohort, respectively (**Figures 1b** and **1d**).

**Figure 1.**
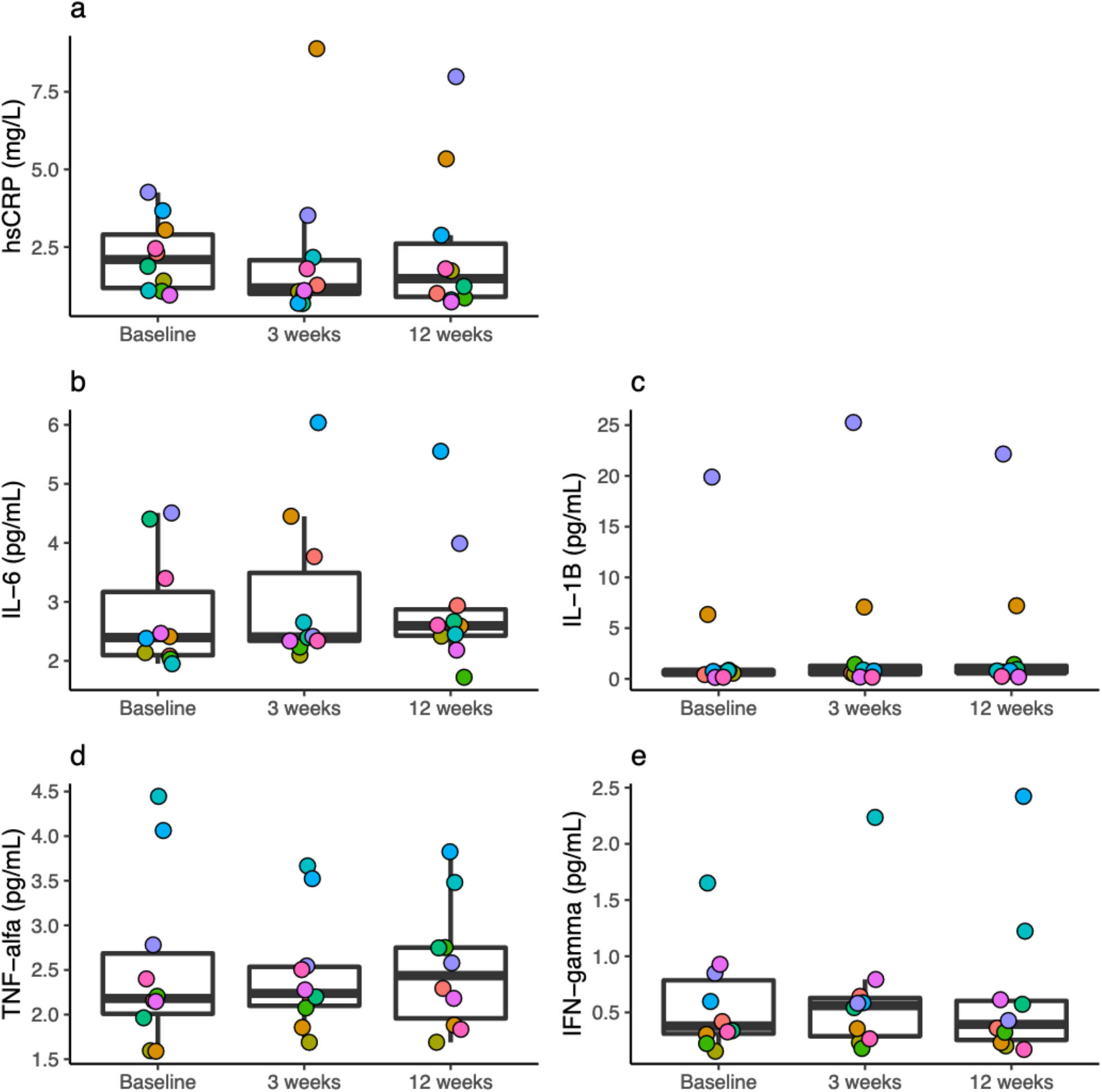
The drug-drug interaction between warfarin and glucose-lowering treatment is associated with the change in glycated hemoglobin (HbA_1c_) during treatment. (**a**) and (**c**) Association between baseline HbA_1c_ and change in INR before and five to seven weeks after initiation of glucose-lowering therapy in the Danish and the Scottish cohorts. (**b**) and (**d**) Association between change in HbA_1c_ from before to two to six months after initiation of glucose-lowering treatment and change in INR before and five to seven weeks after initiation of glucose-lowering therapy in the Danish Cohort and the Scottish cohorts.

### Clinical pharmacokinetic trial

We screened 20 individuals for eligibility and included ten patients with treatment-naïve type 2 diabetes (**Figure S2**). Their median HbA_1c_ was 56 mmol/mol (range 48-102) at inclusion. The median age at inclusion was 61 years (interquartile range (IQR): 59-67 years), all were of white descent, and 90% were male. Two patients were smokers. The median number of concomitant drugs was 2 (IQR: 2-5); most prevalently used were lipid-modifying drugs (60%), thiazides (40%), calcium channel blockers (40%), and ACE inhibitors (40%). Two patients did not receive any concomitant treatment before the study. During the trial, seven patients experienced a total of nine adverse events; four experienced gastrointestinal adverse events related to metformin, two experienced drowsiness associated with midazolam, and three reported unrelated adverse events. One patient started treatment with an SGLT2 inhibitor after the Baseline visit (day 2) due to severe symptomatic hyperglycemia.

All patients were compliant, with metformin intake >90% of expected. Complete pharmacokinetic data were obtained for midazolam (CYP3A4) due to the short elimination half-life. Compared to baseline, three and 12 weeks of metformin treatment did not result in clinically or statistically significant changes in midazolam pharmacokinetics (**Figure 2, Figure 3**, and **Table S1**). The metabolic ratio of caffeine (CYP1A2), efavirenz (CYP2B6), losartan (CYP2C9), omeprazole (CYP2C19), metoprolol (CYP2D6) did not reveal any changes following metformin treatment (**Figure 3** and **Table S1**). The patients showed considerable interindividual variation in drug pharmacokinetics (**Figure 3**).

**Figure 2.**
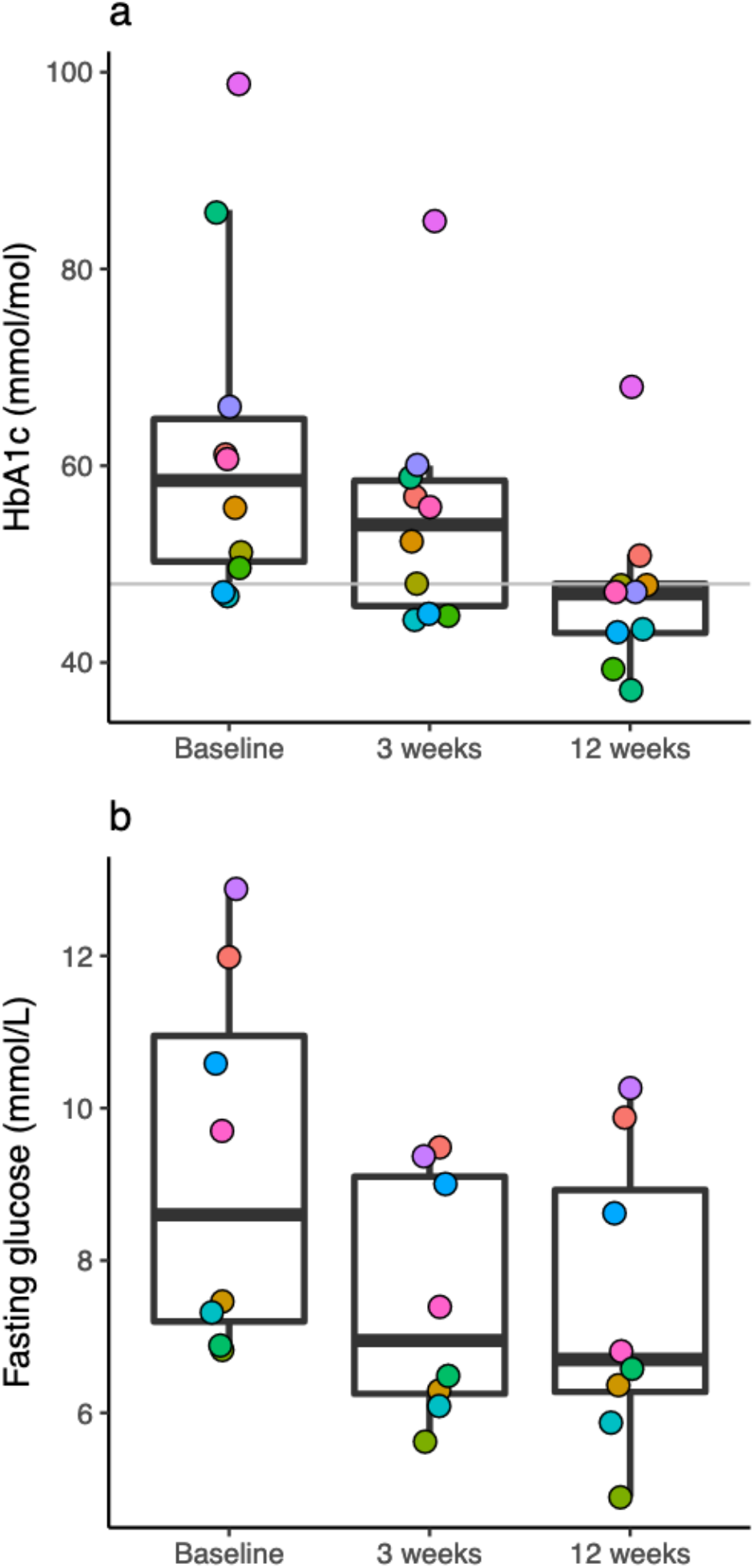
The pharmacokinetics of midazolam (CYP3A4) is unaffected by 3 and 12 weeks of metformin treatment in patients with treatment-naïve type 2 diabetes. The concentration-time curves illustrate mean plasma concentrations. Data is based on nine subjects.

**Figure 3.**
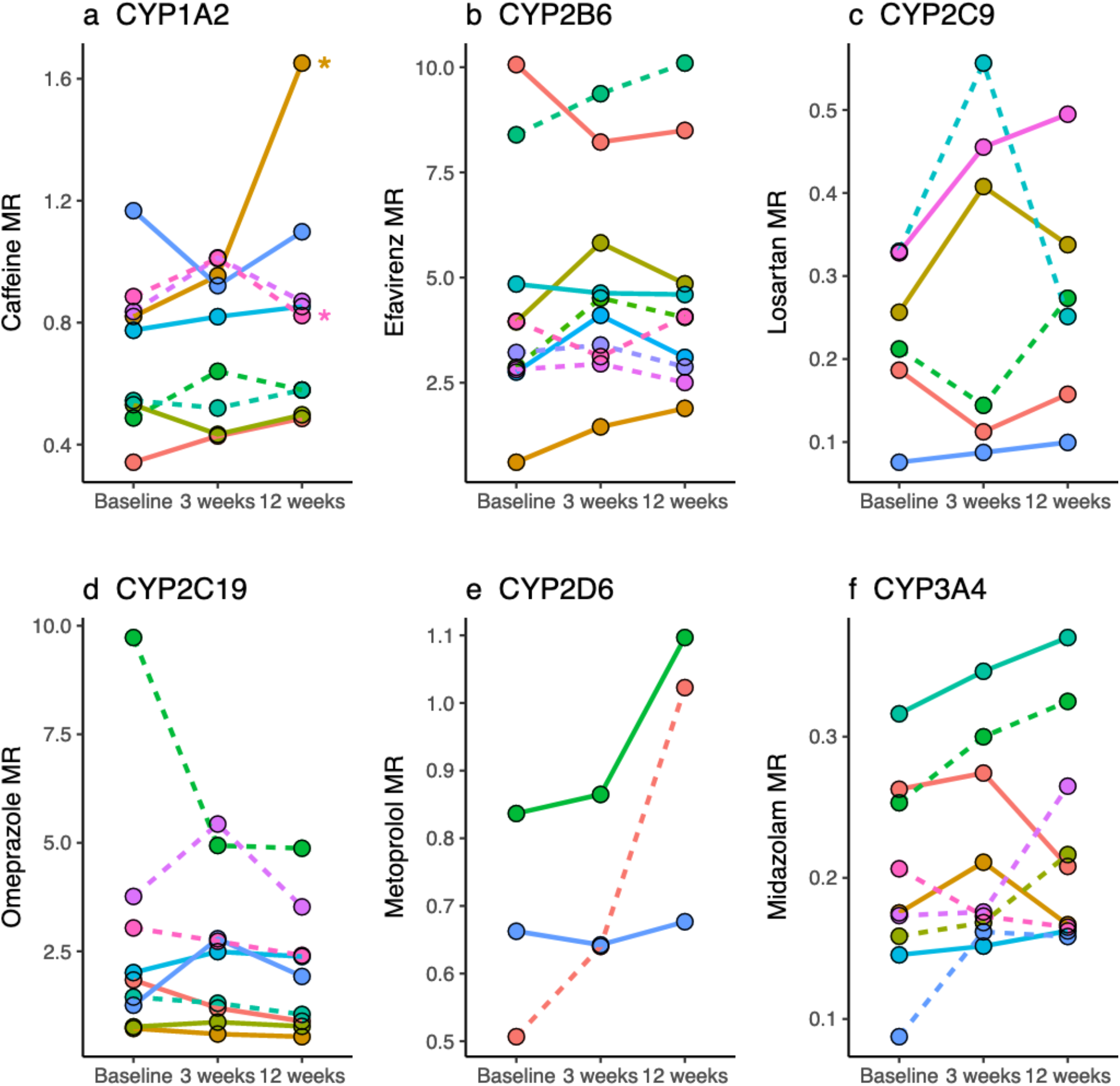
The activity of drug-metabolizing enzymes is unaffected by 3 and 12 weeks of metformin treatment in patients with treatment-naïve type 2 diabetes. Individual metabolic ratios (MR) for each of the six probe drugs are illustrated, and each patient is illustrated with the same color as in **Figure 4** and **Figure 5**. Patients with a large decrease in HbA_1c_ (>-10 mmol/mol) after 12 weeks are shown as dashed lines. **(a)** Metabolic ratio [caffeine]/[paraxanthine] at 4 hours (n=9). * indicates the two patients who were smoking before and during the trial. **(b)** Metabolic ratio [efavirenz]/[8-hydroxyefavirenz] at 6 hours. **(c)** Metabolic ratio ([losartan]/[E3174] at 6 hours (n=6). **(d)** Metabolic ratio [omeprazole]/[hydroxyomeprazole] at 4 hours (n=9). **(e)** Metabolic ratio [metoprolol]/[5-hydroxymetoprolol] at 6 hours (n=3). **(f)** Metabolic ratio [midazolam]/[α-hydroxymidazolam at 2 hours (n=9). Abbreviations: CYP, cytochrome P450 enzymes; MR, metabolic ratio.

All patients had detectable efavirenz plasma concentrations in the sample for 0 hours at 3 weeks, approximately 21 days after the first trial day (baseline). We included this sample in a post-hoc non-compartmental analysis for efavirenz at baseline and determined a median elimination half-life of 96.87 (IQR: 86.11-106.32) for efavirenz (r^2^ > 0.98).

Following genotyping, one patient was identified as a CYP2C9 poor metabolizer, and one patient was identified as a CYP2D6 ultrarapid metabolizer. None of these patients were included in the data analysis for other reasons (**Table S1**). Besides, the patients were found to have normal metabolic activity of CYP2C9, CYP2C19, and CYP2D6.

To assess if a reduction in HbA_1c_ impacts the change in drug metabolism, we applied the same ranges for change in HbA_1c_ as in the register-based study. The magnitude of the change in HbA_1c_ at 12 weeks compared to baseline (low (<10 mmol/mol) vs. high (>10 mmol/mol)) did not affect the metabolic ratios of any of the six probe drugs (p>0.05 for all six probe drugs) **(Figure 3)**.

Bodyweight and BMI were reduced during the trial and were significantly lower at the follow-up after 12 weeks of metformin treatment (**Table 2**). Two patients (20%) obtained a clinically relevant weight loss of 5% or more. Similarly, the median waist circumference was lower after 12 weeks of metformin treatment (**Table 2**).

**Table 2.**
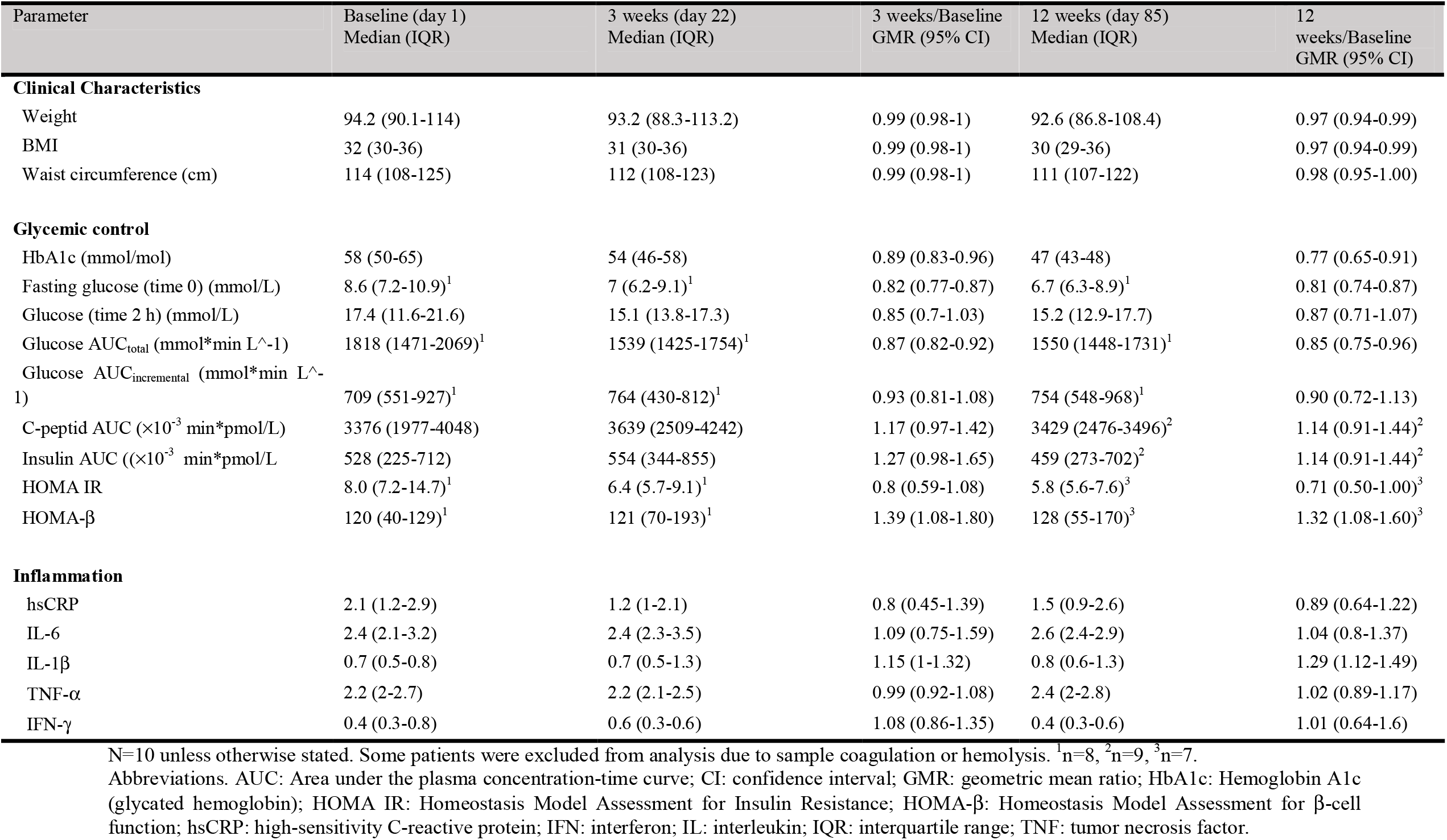
Clinical characteristics and glycemic control improved in 10 patients with treatment-naïve type 2 diabetes following three weeks and 12 weeks of metformin treatment compared to baseline. Inflammation remained low and unchanged during the course of metformin treatment.

All patients improved their glycemic control after 3 and 12 weeks of metformin treatment (**Table 2**). The median change in HbA_1c_ after 3 and 12 weeks of metformin treatment was -4 mmol/mol (IQR: -6;-3 mmol/mol) and -10 mmol/mol (IQR: -18;-5 mmol/mol), respectively). After 12 weeks, 5 patients (50%) had obtained a reduction in HbA_1c_ >10 mmol/mol. At the end of the trial, 60% of the patients reached the therapeutic goal of HbA_1c_ <48 mmol/mol (**Figure 4)**. The total exposure to glucose reduced following metformin treatment; fasting glucose and glucose AUC_0-2 h_ was significantly lower after three weeks and 12 weeks of treatment than baseline. However, mean fasting glucose at 2 hours remained unchanged at 11 mmol/L. Surrogate measures of insulin sensitivity and secretion improved in response to metformin treatment and glycemic control (**Table 2**).

**Figure 4.**
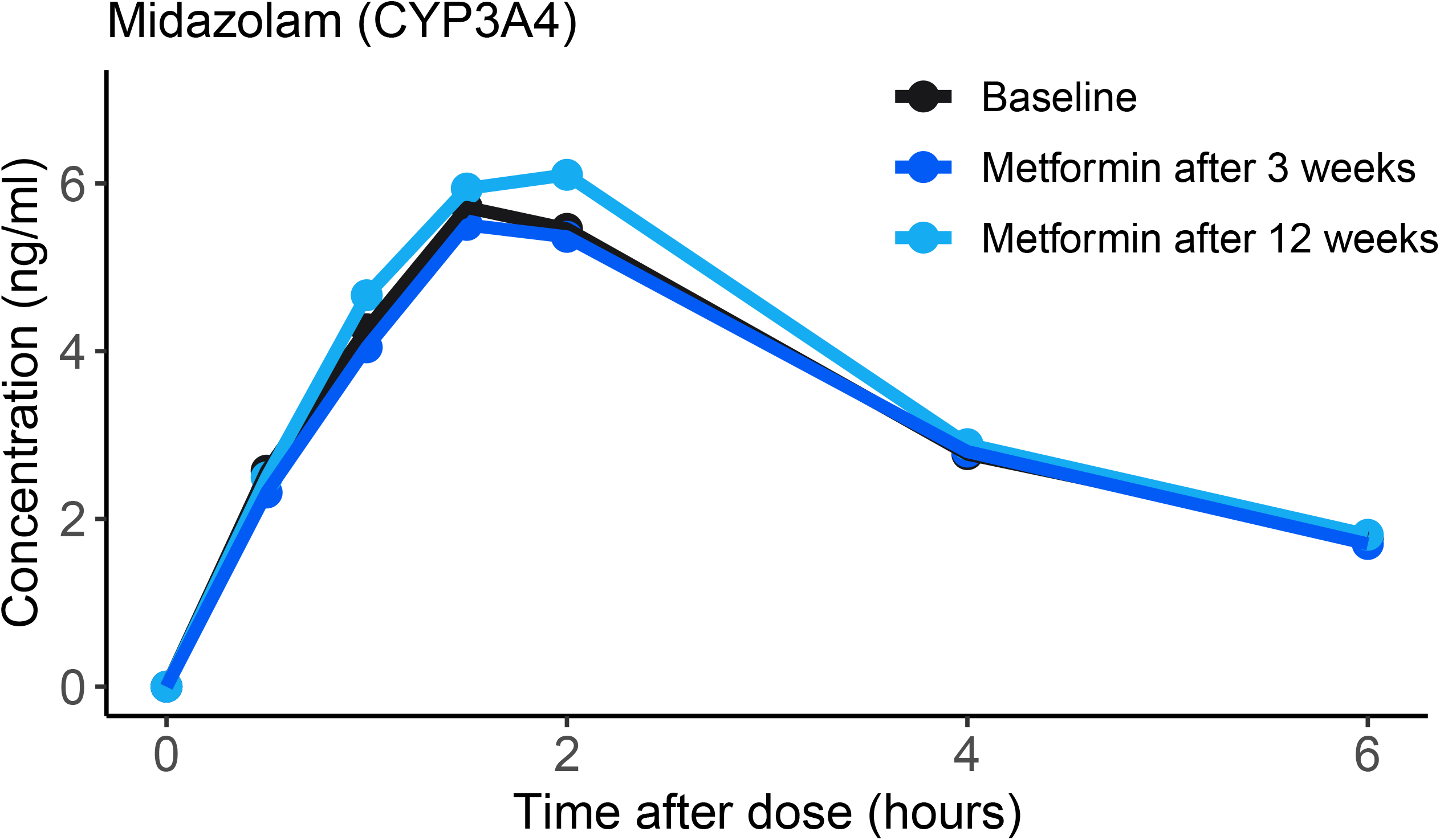
All 10 treatment-naïve patients with type 2 diabetes responded to metformin treatment assessed by glycated hemoglobin (HbA_1c_) (**a**) and fasting glucose (**b**) after 3 and 12 weeks of treatment with metformin, compared to baseline. Boxes are median and interquartile ranges. Each patient is illustrated with the same color as in **Figure 3** and **Figure 5**. Two patients had incomplete sampling due to sample coagulation and were not shown (n=8). **(a)** HbA_1c_ (mmol/mol), the grey horizontal line marks 48 mmol/mol, the diagnostic limit and standard therapeutic goal of type 2 diabetes. Note that two patients had a slight decrease in HbA_1c_ from inclusion (inclusion criteria HbA_1c_ ≥48 mmol/mol) to the Baseline visit **(b)** Fasting plasma glucose (mmol/L), time 0.

All patients had detectable levels of the inflammatory marker hsCRP and the pro-inflammatory cytokines IL-6, IL-1β, TNF-α, and IFN-γ. IL-1β increased slightly after 12 weeks of metformin treatment, but levels remained at a clinically insignificant low level. Throughout the trial, the levels of hsCRP, IL-6, TNF-α, and IFN-γ remained low and unchanged following metformin treatment and improved glycemic control (**Table 2** and **Figure 5**).

**Figure 5.**
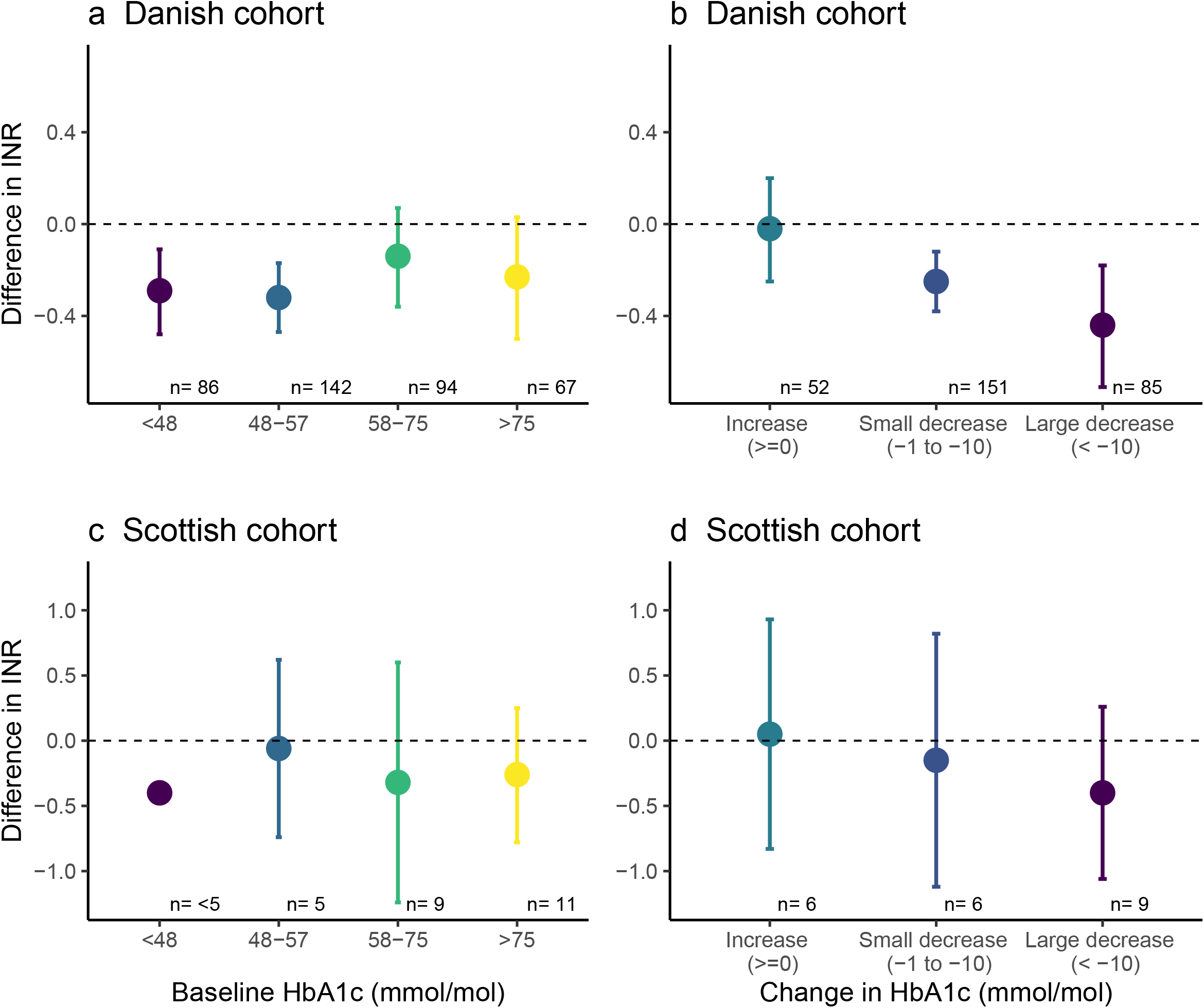
Inflammation remained low and unchanged in 10 patients with treatment-naïve type 2 diabetes following three and 12 weeks of metformin treatment compared to baseline. Boxes mark the median and interquartile ranges. Each patient is illustrated with the same color as in **Figure 3** and **Figure 4. (a)** High-sensitivity C-Reactive Protein (hsCRP) (mg/L). **(b)** Interleukin 6 (IL-6) (pg/mL). (**c**) Interleukin 1beta (IL-1β) (pg/mL). (**d**) Tumor Necrosis Factor alfa (TNF-α) (pg/mL). (**e**) Interferon-gamma (IFN-γ) (pg/mL).

## DISCUSSION

This translational study demonstrates that chronic warfarin users risk decreased warfarin efficacy when initiating glucose-lowering treatment in a register-based study. Furthermore, we show that this is not caused by altered drug metabolism in a clinical pharmacokinetic trial. Initiating metformin in patients with treatment-naïve type 2 diabetes did not affect the activity of drug-metabolizing enzymes (CYP1A2, CYP2B6, CYP2C9, CYP2C19, CYP2D6, and CYP3A4) following three and 12 weeks of metformin treatment, compared to baseline. This is despite a marked improvement in glycemic control following metformin treatment. The level of inflammation remains low and unchanged during metformin treatment.

In this study, we confirm the previous findings that warfarin efficacy is reduced following the initiation of glucose-lowering treatment (8,9). Moreover, we demonstrate that patients with a strong effect of glucose-lowering therapy have a more pronounced reduction in warfarin efficacy. Several studies have found lower CYP activity, most importantly CYP3A4, in patients with type 2 diabetes compared to healthy controls (12–14). We speculated that the initiation of glucose-lowering therapy might reverse the diabetes-induced suppression of drug-metabolizing enzymes. As observed in our register-based studies, this would lead to increased warfarin metabolism and reduced therapeutic efficacy. However, our clinical pharmacokinetic trial does not support this hypothesis. From a clinical perspective, the results are reassuring, and the observed effect seems limited to warfarin. Vitamin K is the primary target of warfarin, and glucose-dependent effects on the vitamin K pathway or warfarin sensitivity are potential mechanisms for the observed effect of glucose-lowering drugs. Further exploratory studies are warranted to understand the mechanism leading to decreased warfarin efficacy after initiating glucose-lowering drugs.

This is the first self-controlled clinical pharmacokinetic trial assessing the impact of initiating a glucose-lowering drug on the activity of multiple drug-metabolizing enzymes. Previous studies have focused on the drug-metabolizing activity (12–14) and the content of drug-metabolizing enzymes (14,27) in patients with type 2 diabetes compared to healthy individuals. In line with previous studies (12–14,28), our results suggest that the drug-metabolizing activity of CYP2C19 and CYP3A4 is decreased, and CYP1A2 activity is increased in patients with type 2 diabetes compared to healthy individuals (manuscript in the draft, *ClinicalTrials*.*gov identifier NCT04840641)*. Further studies are needed to assess the mechanism behind this difference.

We observed efavirenz to have an elimination half-life far exceeding the expected (29,30); all patients had detectable plasma concentrations of efavirenz 21 days after taking a single dose. In the product labels, the single dose elimination half-life is reported to be 52-76 hours (29,30), and 21 days should be sufficient to clear efavirenz after administration. A study assessing the steady-state pharmacokinetics of efavirenz reported an elimination half-life ranging from 27-136 hours following discontinuation of steady-state treatment (31). We find elimination half-lives from 78-147 hours in patients with type 2 diabetes; this estimate might even be too low as efavirenz elimination is biphasic, and more data points are required to determine the terminal elimination. Based on the observation of prolonged elimination half-life among patients with type 2 diabetes, we hypothesize that the mechanisms underlying this potential impactful regulation of CYP2B6 and efavirenz metabolism may be caused by the pathophysiological properties of type 2 diabetes. Further studies are warranted to address this.

The main strength of this study is the translational approach. The register-based study allows us to evaluate the impact on a population level, while the clinical pharmacokinetic trial provides mechanistic insight into this putative drug-drug interaction. We replicated the population-level data in two geographically distinct cohorts with many chronic warfarin users. Using a self-controlled design eliminates interindividual variability and provides accurate estimates. The primary limitation is the lack of clinical endpoints related to decreased INR, e.g., death and thromboembolic events. Furthermore, we interpreted redeeming a prescription as the initiation of a glucose-lowering drug. Thus, ingestion of the medication is not guaranteed. HbA_1c_ data suggest that some individuals are non-responsive, non-compliant, or had an increase in HbA_1c_ in the time window between the last HbA_1c_ and the index date. Exposure misclassification of these individuals, with little or no change in INR, might underestimate the impact on warfarin efficacy.

The design of the clinical trial includes several strengths. We included patients with a medical indication for treatment with metformin, making the results generalizable to other patients initiating glucose-lowering drugs. We included males and females with a broad range of ages and comorbidities based on concomitant medication. The significant heterogeneity among trial subjects is accounted for in the self-controlled design, leaving the estimates unaffected by interindividual differences. Lastly, we have limited the sampling time to 6 hours, ensuring the inclusion of a representative proportion of treatment-naïve patients with type 2 diabetes and avoiding a potential socioeconomic bias in the patients included. However, the shorter sampling time is also the main limitation of the clinical pharmacokinetic trial. Complete pharmacokinetics of all six probe drugs would require 72 hours of sampling and allow assessment of all pharmacokinetic parameters rather than metabolic ratios. The metabolic ratios were identified in a cohort of healthy volunteers (19,26), but it is unknown if metabolic ratios correlate to drug exposure in patients with type 2 diabetes. Another limitation of the trial is the lack of metformin concentrations in the trial subjects. Thus, metformin exerts some of its glucose-lowering effects in the intestine (6), and the correlation between metformin exposure and response is limited (32). Lastly, we observed inflammation to be low and unchanged after metformin treatment. However, inflammation might be different in patients with longer diabetes duration, which could potentially impact CYP activity. We did not require the trial subjects to have a reduction in blood glucose during the trial, which could maximize the observed impact. Introducing such restrictions in the trial design would decrease the number of trial subjects available for analysis. We do not observe the reduction in HbA_1c_ to affect the drug-metabolizing activity.

In conclusion, initiation of glucose-lowering drugs in patients with chronic warfarin use is associated with a reduced anticoagulant efficacy of warfarin that needs special clinical attention to avoid thrombotic complications. We show that this is not caused by altered drug metabolism in treatment-naïve patients with type 2 diabetes that initiates glucose-lowering therapy. Further studies are needed to understand whether the reduction in warfarin efficacy leads to an increased risk of clinical endpoints such as stroke and death. Finally, additional work is required to understand the underlying mechanisms of this effect.

## STUDY HIGHLIGHTS

### What is the current knowledge on the topic?

- Patients with type 2 diabetes might have lower activity of drug-metabolizing enzymes compared to healthy controls.
- Glucose-lowering drugs might reverse this suppression and normalize drug metabolism and have been shown to reduce the anticoagulant efficacy of warfarin in register-based studies.

### What question did this study address?

- Does initiation of glucose-lowering drugs affect the anticoagulant efficacy of warfarin and drug metabolism in patients with type 2 diabetes?

### What does this study add to our current knowledge?

- Anticoagulant efficacy of warfarin decreases after initiation of glucose-lowering drugs, particularly among individuals with a substantial glycemic response in register-based studies.
- Drug metabolism is not affected after initiation of metformin among patients with treatment-naïve type 2 diabetes in a clinical pharmacokinetic trial.

### How might this change clinical pharmacology or translational science?

- Chronic warfarin users have increased risk of reduced warfarin efficacy after initiating glucose-lowering drugs. The effect appears unrelated to drug metabolism and might be isolated to warfarin. Further studies should focus on the mechanisms underlying altered drug metabolism in patients with type 2 diabetes compared to healthy controls.

## Data Availability

Data cannot be made publicly available.

## Acknowledgment

We acknowledge the support of the Health Informatics Centre, University of Dundee, for managing and supplying the anonymized data for the Scottish cohort. We thank the general practices for recruiting patients with type 2 diabetes and the patients for providing their time for the trial. We thank Birgitte Damby Sørensen, Rasmus Andersen, Anette Tyrsted, Lone Hansen, Charlotte Bøtchiær Fage Olsen, Sara Egsgaard, and Camilla Davidsen for their hard work in planning and performing biochemical analysis. Lastly, we thank OPEN, Open Patient data Explorative Network, Odense University Hospital, Region of Southern Denmark, for support and facilities for hosting the clinical trial data and case report form (CRF).

All statistical analysis is conducted with Stata 15.1 (StataCorp. 2017. Stata Statistical Software: Release 15. College Station, TX: StataCorp LLC) and RStudio Version 1.3.1056 (RStudio Team (2020). RStudio: Integrated Development for R. RStudio, PBC, Boston, MA). Figure 1 is created with Biorender.com.

## Author contributions

Ann-Cathrine Dalgård Dunvald, Anton Pottegård, and Tore B. Stage designed the research. All authors participated in obtaining data sources or conception and design of the studies. Ann-Cathrine Dalgård Dunvald performed the research. Flemming Nielsen, Dorte Aalund Olsen, and Jonna Skov Madsen contributed new reagents/analytical tools. Ann-Cathrine Dalgård Dunvald, Martin Thomsen Ernst, Louise Donnelly, and Enrique Soto-Pedre conducted the data analysis. All authors revised the manuscript critically for important intellectual content, and all authors have read and approved the final version to be published.

