## Supplementary for "Initiation of glucose-lowering drugs reduces the anticoagulant effect of warfarin – but not through altered drug metabolism in patients with type 2 diabetes"

1. Clinical Pharmacology, Pharmacy, and Environmental Medicine. Department of Public Health, University of Southern Denmark, Denmark
2. Department of Biochemistry and Immunology, Lillebaelt Hospital, University Hospital of Southern Denmark, Vejle, Denmark
3. Department of Population Health & Genomics, School of Medicine, University of Dundee, Scotland, UK
4. Steno Diabetes Center Odense (SDCO), Odense University Hospital , Odense, Denmark
5. Department of Clinical Research, University of Southern Denmark, Denmark
6. Steno Diabetes Center Copenhagen, Herlev Hospital, Herlev, Denmark
7. Department of Regional Health Research, Faculty of Health Sciences, University of Southern Denmark, Denmark
8. Research Unit of General Practice, Department of Public Health, University of Southern Denmark, Denmark
9. Department of Clinical Pharmacology, Odense University Hospital, Odense, Denmark

**Correspondance**
Tore B. Stage
Clinical Pharmacology, Pharmacy and Environmental Medicine
University of Southern Denmark
JB Winsløws Vej 19, 2
DK-5000 Odense C, Denmark


### Methods

#### Clinical pharmacokinetic trial

*Basel cocktail:*

- 100 mg caffeine (Cofi-Tabs (dietary supplement), Vitabalans, Finland))
- 50 mg efavirenz (Stocrin®, MSD, Denmark)
- 12.5 mg losartan (Losartan "Medical Valley," Medical Valley, Sweden)
- 10 mg omeprazole (Omeprazole "Medical Valley," Medical Valley, Sweden)
- 12.5 mg metoprolol (Metoprololsuccinat "Hexal," Sandoz, Denmark)
- 2 mg midazolam (oral solution, Ozalin, Primex Pharmaceuticals, Finland))

*Sampling on trial days*

An intravenous catheter was placed in the antecubital vein for blood sampling. Blood samples were collected in K2-EDTA-, Na-Florid-Citrate-, and gel-containing tubes at the following time points: 0 (before administration of the Basel cocktail), 0.5, 1, 1.5, 2, 4 (only EDTA), and 6 hours (only EDTA) (**Figure S1**). Samples for plasma were centrifuged at 2500 g for 10 minutes within 2 hours, and plasma was transferred into cryo-tubes for storage at -20°C and -80°C. Samples for serum were coagulated for at least 30 minutes before centrifuging at 2100 g for 10 minutes at 8°C, and serum was transferred into cryo-tubes for storage at -80°C. The participants collected urine in intervals of 0 to 6 hours. After mixture, urine was transferred into cryotubes for storage at -20°C.

*Analytical methods*

The following drugs and metabolites were analyzed in EDTA plasma and urine in the clinical pharmacokinetic trial: caffeine, paraxanthine, efavirenz, 8-hydroxyefavirenz, losartan, E3174, omeprazole, 5-hydroxyomeprazole, metoprolol, hydroxymetoprolol, midazolam, and α-hydroxymidazolam.

We used high-performance liquid chromatography (LC) and high-resolution mass spectrometry (HR-MS) for the drug and metabolite concentrations. The LC-HR-MS system consisted of a Thermo Vanquish UHPLC system equipped with a split sampler FT, a binary pump F, and a column compartment connected to a Q-Exactive Orbitrap high-resolution mass spectrometer with a heated electrospray ionization (H-ESI) interface (Thermo Fisher Scientific Inc., Waltham, MA). The analytical column utilized was an ACE 3 C18-AR 50 x 2.1 mm with 3 µm particle size

(Advanced Chromatography Technologies Ltd., Aberdeen, UK), operated at a column temperature of 40°C.

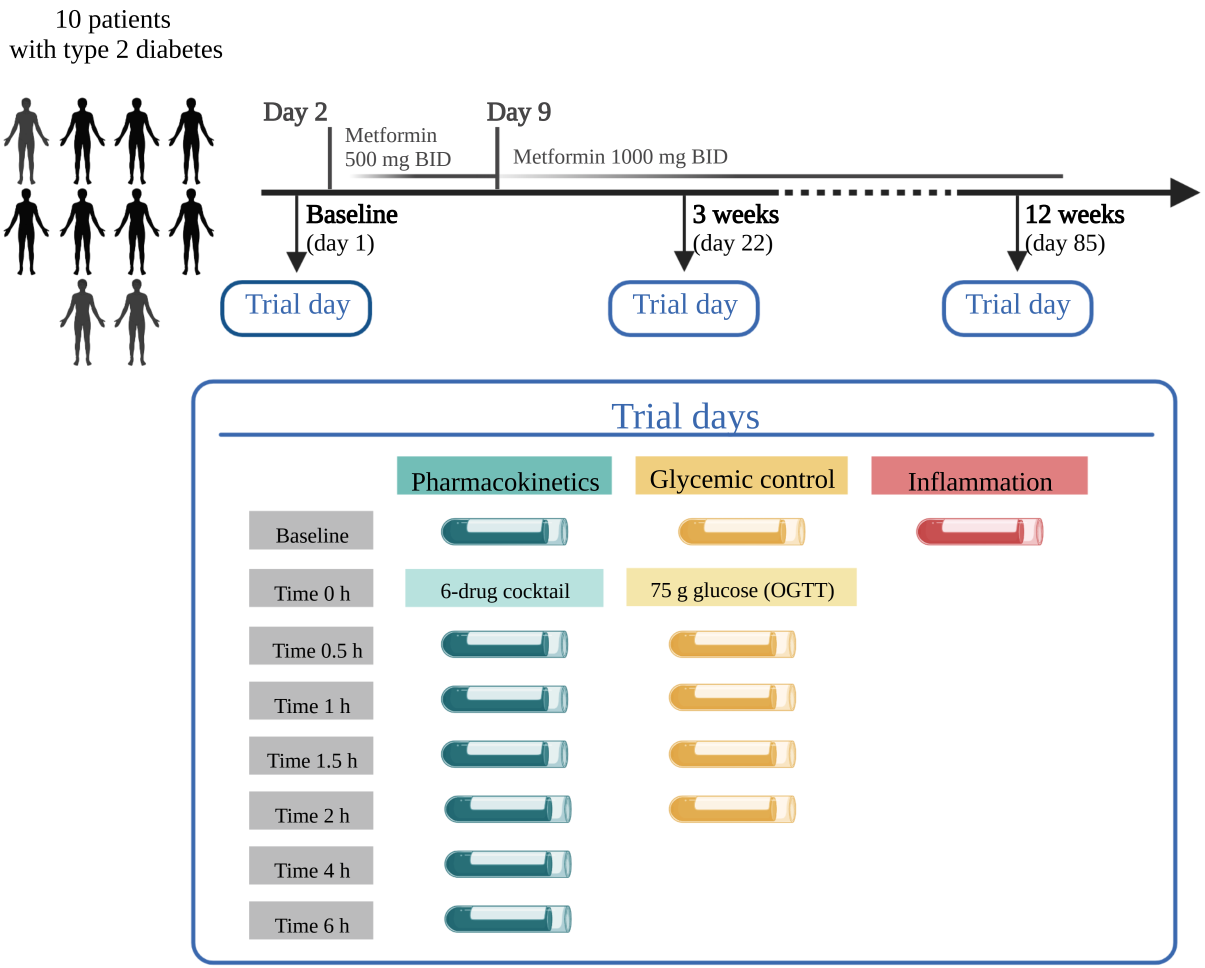


**Figure S1** Schematic overview of the trial design and sampling schedule for this self-controlled, clinical pharmacokinetics trial.
Abbreviations: BID: bis in die (twice a day), h: hour, OGTT: oral glucose tolerance test

**Figure S2** Flow diagram of the screening process. Twenty individuals were screened according to inclusion and exclusion criteria, of which ten patients with type 2 diabetes were eligible for inclusion in this clinical pharmacokinetic trial.

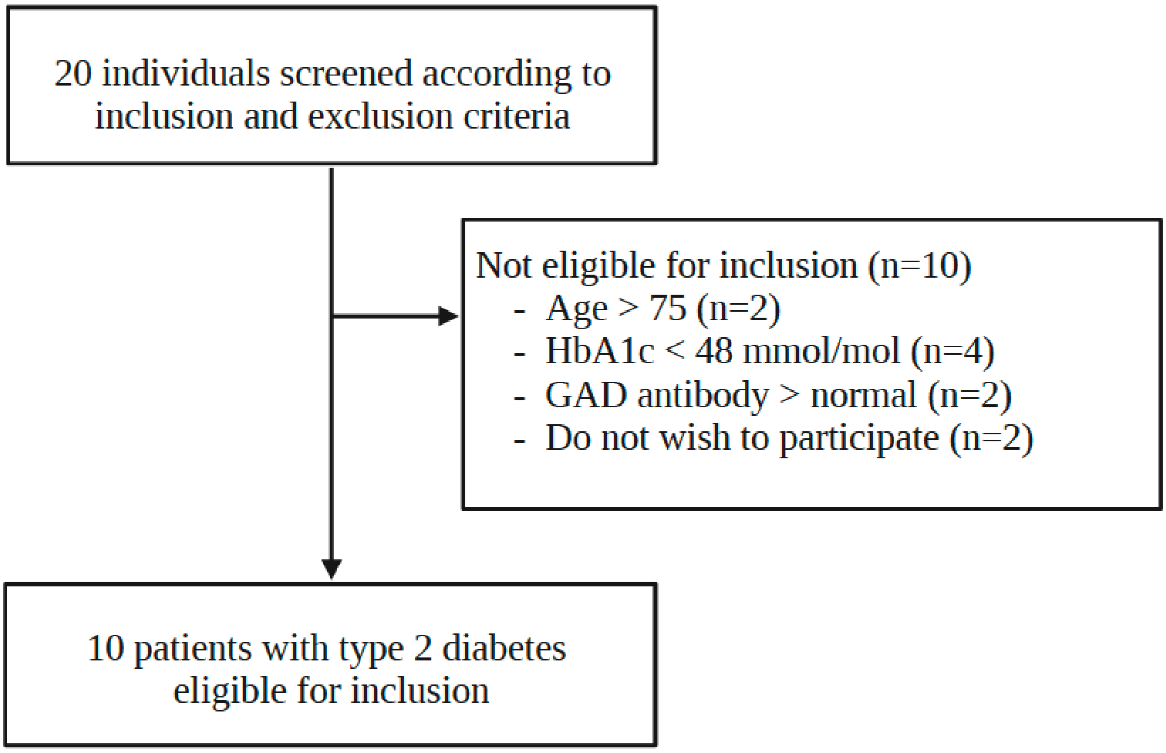


Abbreviations: GAD, Glutamic Acid Decarboxylase 65; n, number.

**Supplementary Table S1** Pharmacokinetic parameters and metabolic ratios of the probe drugs caffeine (CYP1A2), efavirenz (CYP2B6), losartan (CYP2C9), omeprazole (CYP2C19), metoprolol (CYP2D6), midazolam (CYP3A4) were unaffected by 3 and 12 weeks of metformin treatment. Assessed in 10 patients with treatment-naïve type 2 diabetes at Baseline (day 1) and after 3 weeks (day 22) and 12 weeks (day 85) of metformin treatment.

| | **Baseline** (day 1) Median (IQR) | **3 weeks** (day 22) Median (IQR) | **3 weeks/Baseline** GMR (95% CI) | **12 weeks** (day 85) Median (IQR) | **12 weeks/Baseline** GMR (95% CI) |
|---|---|---|---|---|---|
| CAFFEINE (CYP1A2)[1] | | | | | |
| Cmax (ng ml^-1) | 1134 (1025-1743) | 1151 (963-1325) | 0.91 (0.81-1.02) | 1078 (1017-1322) | 0.92 (0.84-1.01) |
| Tmax (h) | 2 (2-4) | 2 (2-4) | 0.92 (0.77-1.11) | 2 (2-2) | 0.92 (0.77-1.11) |
| Metabolic ratio [par]/[caf][b] | 0.77 (0.53-0.84) | 0.82 (0.52-0.95) | 1.06 (0.92-1.22) | 0.82 (0.58-0.87) | 1.15 (0.94-1.39) |
| Metabolic ratio [caf]/[par][b] | 1.29 (1.2-1.88) | 1.22 (1.05-1.92) | 0.94 (0.82-1.08) | 1.21 (1.15-1.73) | 0.87 (0.72-1.06) |
| EFAVIRENZ (CYP2B6) | | | | | |
| Cmax (ng ml^-1) | 185 (159-213) | 199 (174-246) | 1.11 (0.95-1.29) | 194 (141-235) | 1.01 (0.83-1.21) |
| Tmax (h) | 4 (4-4) | 4 (4-4) | 1.00 (0.99-1.01) | 4 (4-4) | 0.91 (0.70-1.18) |
| Metabolic ratio [OH-efa]/[efa][c] | 0.28 (0.22-0.35) | 0.23 (0.18-0.31) | 0.83 (0.65-1.06) | 0.25 (0.21-0.34) | 0.86 (0.65-1.12) |
| Metabolic ratio [efa]/[OH-efa][c] | 3.58 (2.83-4.62) | 4.31 (3.19-5.53) | 1.2 (0.94-1.53) | 4.06 (2.93-4.79) | 1.17 (0.89-1.54) |
| LOSARTAN (CYP2C9)[2] | | | | | |
| Cmax (ng ml^-1) | 15.3 (11.6-21.1) | 11.7 (10.4-17.6) | 0.86 (0.74-1.00) | 13.8 (11.2-19.2) | 0.93 (0.70-1.24) |
| Tmax (h) | 4 (2.5-4) | 4 (4-4) | 1.25 (0.86-1.83) | 4 (4-4) | 1.26 (0.86-1.85) |
| Metabolic ratio [E3174]/[los][c] | 4.31 (3.26-5.2) | 4.69 (2.26-8.41) | 0.91 (0.57-1.44) | 3.82 (3.14-5.75) | 0.88 (0.66-1.18) |
| Metabolic ratio [los]/[E3174][c] | 0.23 (0.19-0.31) | 0.28 (0.12-0.44) | 1.1 (0.69-1.74) | 0.26 (0.18-0.32) | 1.14 (0.85-1.52) |
| OMEPRAZOLE (CYP2C19)[3] | | | | | |
| Cmax (ng ml^-1) | 95.5 (67.5-157.1) | 112.1 (87.8-159.8) | 1.12 (0.82-1.54) | 125. 8 (74.2-177.4) | 1.17 (0.82-1.66) |
| Tmax (h) | 4 (4-4) | 4 (4-4) | 0.92 (0.82-1.04) | 4 (4-4) | 0.96 (0.88-1.05) |
| Metabolic ratio [OH-ome]/[ome][b] | 0.54 (0.33-0.80) | 0.40 (0.36-0.84) | 1.00 (0.71-1.4) | 0.52 (0.42-1.12) | 1.21 (0.91-1.62) |
| Metabolic ratio [ome]/[OH-ome][b] | 1.84 (1.26-3.04) | 2.49 (1.19-2.79) | 1 (0.71-1.4) | 1.92 (0.89-2.41) | 0.82 (0.62-1.1) |
| METOPROLOL (CYP2D6)[4] | | | | | |
| Cmax (ng ml^-1) | 1.3 (1.0-1.8) | 1.2 (1.2-1.7) | 1.02 (0.68-1.54) | 1.8 (1.3-1.8) | Inf (NaN-NaN) |
| Tmax (h) | 6 (4.5-6) | 6 (6-6) | 1.07 (0.78-1.47) | 6 (6-6) | 1.14 (0.92-1.42) |
| Metabolic ratio [OH-met]/[met][c] | 1.51 (1.35-1.74) | 1.56 (1.36-1.56) | 0.92 (0.66-1.3) | 0.98 (0.94-1.23) | 0.72 (0.3-1.69) |
| Metabolic ratio [met]/[OH-met][c] | 0.66 (0.58-0.75) | 0.64 (0.64-0.75) | 1.08 (0.77-1.52) | 1.02 (0.85-1.06) | 1.39 (0.59-3.28) |
| MIDAZOLAM (CYP3A4)[5] | | | | | |
| $AUC_{0\text{-}6h}$ (ng*h ml^-1) | 18.3 (13.9-23.7) | 18.0 (14.4-23.7) | 0.99 (0.90-1.09) | 18.6 (16.8-23.6) | 1.1 (0.98-1.25) |
| AUC 0-Inf (ng*h ml^-1) | 24.0 (16.8-30.4) | 22.2 (17.6-30.7) | 0.93 (0.81-1.08) | 22.8 (20.7-35.9) | 1.05 (0.90-1.22) |
| Cmax (ng ml^-1) | 5.62 (4.67-5.89) | 5.66 (4.37-6.93) | 1.00 (0.87-1.15) | 6.15 (5.27-6.43) | 1.15 (1.01-1.31) |
| Tmax (h) | 2 (1.5-2) | 2 (2-2) | 1.06 (0.78-1.44) | 1.5 (1.5-2) | 0.93 (0.77-1.12) |
| T1/2 (h) | 2.29 (1.99-2.73) | 2.22 (1.99-2.36) | 0.87 (0.64-1.18) | 2.16 (2.1-2.35) | 0.90 (0.76-1.05) |
| CL/F (L h^-1) | 501 (395-605) | 540 (391-589) | 1.10 (0.97-1.24) | 527 (335-579) | 1.00 (0.88-1.13) |
| Metabolic ratio [OH-mid]/[mid][a] | 5.71 (3.95-6.3) | 5.69 (3.65-5.94) | 0.89 (0.76-1.05) | 4.81 (3.77-6.06) | 0.86 (0.69-1.07) |
| Metabolic ratio [mid]/[OH-mid][a] | 0.18 (0.16-0.25) | 0.18 (0.17-0.27) | 1.12 (0.95-1.32) | 0.21 (0.17-0.26) | 1.16 (0.93-1.44) |

[a] Metabolic ratio is assessed at 2 hours. [b] Metabolic ratio is assessed at 4 hours. [c] Metabolic ratio is assessed at 6 hours.

[1]1 patient consumed caffeine prior to the trial day and was excluded from the analysis (n=9), [2]4 patients had a chronic intake of losartan and concentrations above the limit of detection at baseline and were excluded from the analysis (n=6), [3]1 patient had metabolite concentrations below the limit of detection and was excluded from analysis (n=9) [4]7 patients had plasma concentrations below the limit of detection (drug and/or metabolite) at 6 hours and were excluded from the analysis (n=3), [5]1 patient had incorrect dosing of midazolam at Baseline and was excluded from the analysis (n=9).

Abbreviations: caf: caffeine, par: paraxanthine, efa: efavirenz, OH-efa: 8-hydroxyefavirenz, los: losartan, ome: omeprazole, OH-ome: hydroxyomeprazole, met: metoprolol, OH-met: 5-hydroxymetoprolol, mid: midazolam, OH-mid: α-hydroxymidazolam.
